# Primary Care Practitioners’ Response to 2019 Novel Coronavirus Outbreak in China

**DOI:** 10.1101/2020.02.11.20022095

**Authors:** Zhijie Xu, Yi Qian, Lizheng Fang, Mi Yao

**Author notes:** Corresponding Author: Mi Yao, MD, Institute of Applied Health Research, University of Birmingham, Edgbaston Birmingham, B15 2TT, UK).

## Abstract

The emerging outbreak of the 2019 novel coronavirus (2019-nCoV) originated from Wuhan poses a great challenge to healthcare system in China.^1^ Primary care practitioners (PCPs) have an important role in district communicable disease control.^2^ However, because primary health-care system in China still needs to be substantially strengthened,^3,4^ whether PCPs are proactive and capable in responding to the outbreak remains unclear. Using an electronic questionnaire, we surveyed a national sample of PCPs to assess their response to novel coronavirus outbreak.

## Methods

From Jan 28, 2020 to Feb 3, 2020, we invited a national random sample of PCPs in China to participate in an online survey through social media. The questionnaire was adapted from The Notice of Improving Prevention of 2019-nCoV causing Pneumonia in Primary Care Settings issued by China’s National Health Commission on Jan 27, 2020.^5^ It included 6 questions of demographic information, duration and scope of clinical practice as well as types of practice settings, and 8 statements regarding whether the PCPs were making efforts to respond to novel coronavirus outbreak in daily practice.

Eligible PCPs participated in this survey voluntarily and were not compensated. Once completed and returned, the survey was regarded as informed consent by participants. This study was deemed exempt from review by the Sir Run Run Shaw Hospital Ethics Committee.

One of the authors (Z. X.) reviewed the returned questionnaires and excluded the unqualified data. All statistical analyses were performed with Stata version 15.0 and a two-sided P_<_.05 were considered statistically significant.

## Results

Among 1751 respondents (response rate: 62%), 784 (45%) were men and 1135 (65%) were family physicians. The mean (SD) age of respondents was 38(8) years, and the duration of practice was 15 (9) years (**Table 1**). Most of PCPs were proactive in studying knowledge and skills of containing 2019-nCoV (99%, n=1741), teaching their consulting patients ways of prevention (94%, n=1650), and comforting the worried patients (86%, n=1509). The study also showed that 69% (n=1221) of PCPs used online consultation for health education. However, less than half (48%, n=849) reported to transfer the suspected infected patients to hospitals for further diagnosis and treatment. The rates of visiting, transferring and monitoring the suspected cases varied among PCPs in different practice settings, geographic regions, and specialties, respectively (**Table 2**).

**Table1.** Characteristics of Respondents

| Characteristics | Finding<br>(N=1751) |
| --- | --- |
| Age, mean (SD), y | 38(8) |
| Male sex, No.(%) | 784 (45) |
| Years of practice, y | 15(9) |
| Region,<sup>a</sup> No.(%) |  |
| Northeast | 217(12) |
| Center | 256(15) |
| East | 653(37) |
| West | 625(36) |
| Practice setting, No.(%) |  |
| Community health center | 913(52) |
| Township health center | 630(36) |
| Private clinic | 65(4) |
| Village clinic | 143(8) |
| Specialty, No.(%) |  |
| Family medicine | 1135(65) |
| Internal medicine | 286(16) |
| Surgery | 57(3) |
| Traditional Chinese medicine | 110(6) |
| Others<sup>b</sup> | 163(9) |

<sup>a</sup>The division is based on regional socioeconomic status issued by China's National Bureau of Statistics. <sup>6</sup>
<sup>b</sup>Other clinical types include preventive medicine, rehabilitation and stomatology.

**Table2.** PCPs’ response to 2019 novel coronavirus outbreak

| Statement | Yes,<br>No. (%; 95%CI) | No,<br>No. (%; 95%CI) |
| --- | --- | --- |
| I have studied clinical practice guidelines on containing novel coronavirus outbreak | 1741(99;99-100) | 10(1;0-1) |
| I have participated in primary screening suspected cases in daily practice | 1383(79;77-81) | 368(21;19-23) |
| I have visited residents and/or villagers to screen suspected cases<sup>a</sup> | 1002(57;55-60) | 749(43;40-45) |
| I have taught the consulting patients how to prevent infection of 2019 novel coronavirus | 1650(94;93-95) | 101(6;5-7) |
| I have transferred the suspected cases to hospitals for further diagnosis and treatment<sup>b</sup> | 849(48;46-51) | 902(52;49-54) |
| I have follow up and monitored health conditions of the suspected cases<sup>c</sup> | 1027(59;56-61) | 724(41;39-44) |
| I have comforted the consulting patients if they are worried or panic | 1509(86;85-88) | 242(14;12-15) |
| I have provided consultation to residents and/or villagers through the Internet (e.g., WeChat)<sup>d</sup> | 1221(70;68-72) | 530(30;28-32) |

<sup>a</sup>PCPs in village clinics (68%, n=97) were more proactive in home visit than PCPs in other settings (community health center: 59%, n=541; township health center: 55%, n=346; private clinic: 28%, n=18) (P=0.000).
<sup>b</sup>Transfer were more common among PCPs in central China (58%, n=149) than PCPs in other regions (east: 51%, n=335; west: 47%, n=295; northeast: 32%, n=70)(P=.001).
<sup>c</sup>62% (n=706) of PCPs of family medicine followed up and monitored health conditions of the suspected cases, compared with 52% (n=150) of internal medicine, 47% (n=27) of surgery, 54% (n=60) of traditional Chinese medicine and 52% (n=84) of other clinical types (P=.000).
<sup>d</sup>Men were more likely than women to provide online consultation (75%, n=590 vs
65%, n=631; P=.000).

## Discussion

We found that most of China’s PCPs attached great importance to the national epidemic outbreak and had some preparation of knowledge and skills for the emerging situation. They took multiple control measures instructed by health authorities, and the majority spontaneously provide online consultation to local community, which effectively increased the coverage of health services during a pandemic period and strengthened the relationship between PCPs and the public. Nevertheless, the capacity of transferring suspected cases remained relatively inadequate. Hence, China’s hierarchical medical system for managing communicable disease remains to be improved.

The result that PCPs’ responses varied among practitioners, regions and settings may be attributed to the uneven distribution of healthcare providers and resources across the country. It indicates that policies for containing the coronavirus outbreak in primary care should consider different challenges PCPs encounter and adjust measures to individual conditions.

Our study includes several limitations. First, there is a diversity of control measures among districts and PCPs’ response may be affected by local authorities. Second, PCPs other than clinicians are not included in our study because they are not responsible for screening and transfer.

Policy-makers should take steps to improve PCPs’ capacity and consciousness to ready for rapid response, and strengthen collaboration between PCPs and specialists in hospitals.

## Data Availability

None

## Author Contributions

Dr Yao had full access to all the data in the study and takes responsibility for the integrity of the data and the accuracy of the data analysis. Concept and design: Xu.

Acquisition, analysis, or interpretation of data: All authors.

Drafting of the manuscript: Xu, Yao, Qian.

Critical revision of the manuscript for important intellectual content: All authors.

Supervision: Fang.

